# Population-based whole-genome sequencing with constrained gene analysis identifies predisposing germline variants in children with central nervous system tumors

**DOI:** 10.1101/2022.03.01.22271578

**Authors:** Ulrik Kristoffer Stoltze, Jon Foss-Skiftesvik, Thomas van Overeem Hansen, Anna Byrjalsen, Astrid Sehested, David Scheie, Torben Stamm Mikkelsen, Simon Rasmussen, Mads Bak, Henrik Okkels, Michael Thude Callesen, Jane Skjøth-Rasmussen, Anne-Marie Gerdes, Kjeld Schmiegelow, René Mathiasen, Karin Wadt

## Abstract

**Background:** The underlying cause of central nervous system (CNS) tumors in children is largely unknown. In this nationwide, prospective population-based study we investigate rare germline variants across known and putative CPS genes and genes exhibiting evolutionary intolerance of inactivating alterations in children with CNS tumors.

**Methods:** One hundred and twenty-eight children with CNS tumors underwent whole-genome sequencing of germline DNA. Single nucleotide and structural variants in 315 cancer related genes and 2,986 highly evolutionarily constrained genes were assessed. A systematic pedigree analysis covering 3,543 close relatives was performed.

**Results:** Thirteen patients harbored rare pathogenic variants in nine known CPS genes. The likelihood of carrying pathogenic variants in CPS genes was higher for patients with medulloblastoma than children with other tumors (OR 5.9, CI 1.6-21.2). Metasynchronous CNS tumors were observed exclusively in children harboring pathogenic CPS gene variants (n=2, p=0.01).

In general, known pCPS genes were shown to be significantly more constrained than both genes associated with risk of adult-onset malignancies (p=5e^−4^) and all other genes (p=5e^−17^). Forty-seven patients carried 66 loss-of-functions variants in 60 constrained genes, including eight variants in six known pCPS genes. A deletion in the extremely constrained *EHMT1* gene, formerly somatically linked with sonic hedgehog medulloblastoma, was found in a patient with this tumor.

**Conclusions:** ∽10% of pediatric CNS tumors can be attributed to rare variants in known CPS genes. Analysis of evolutionarily constrained genes may increase our understanding of pediatric cancer susceptibility.

**3 key points:**

- ∽10% of children with CNS tumors carry a pathogenic variant in a known cancer predisposition gene
- Known pediatric-onset cancer predisposition genes show high evolutionary constraint
- Loss-of-function variants in evolutionarily constrained genes may explain additional risk

**Importance of this study:** Although CNS tumors constitute the most common form of solid neoplasms in childhood, our understanding of their underlying causes remains sparse. Predisposition studies often suffer from selection bias, lack of family and clinical data or from being limited to SNVs in established cancer predisposition genes. We report the findings of a prospective, population-based investigation of genetic predisposition to pediatric CNS tumors. Our findings illustrate that 10% of children with CNS tumors harbor a damaging alteration in a known cancer gene, of which the majority (9/13) are loss-of-function alterations. Moreover, we illustrate how recently developed knowledge on evolutionarily loss-of-function intolerant genes may be used to investigate additional pediatric cancer risk and present *EHMT1* as a putative novel predisposition gene for SHH medulloblastoma. Previously undescribed links between variants in known cancer predisposition genes and specific brain tumors are presented and the importance of assessing both SV and SNV is illustrated.

## Introduction

Central nervous system (CNS) tumors are the most common form of solid neoplasms during childhood and the leading cause of cancer-related death among children^1^. While considerable progress has been made in understanding the molecular biology of pediatric brain tumors, their underlying causes are largely unknown.

Large-scale pan childhood cancer studies have identified 7-9% of patients as carrying pathogenic variants in a cancer predisposition syndrome (CPS) gene^2,3^. These studies, however, include partly overlapping cohorts with overrepresentation of cancers with poor clinical outcome potentially resulting in misleading variant estimates compared to population-based approaches. Similar studies on pediatric CNS tumors have found pathogenic germline alterations in up to 35%, with estimates varying greatly depending on tumor type focus, sample selection and study methodology^4–7^. Although some have employed population-based approaches, most studies suffer from either small sample sizes, selection bias towards recurrent/high-grade tumors, restricted tumor type focus or from lack of detailed clinical data and relevant family history. Moreover, to be able to process the vast amounts of data originating from whole-genome and whole-exome sequencing (WGS/WES) much of the existing literature is restricted to cancer gene panels and single nucleotide variants (SNVs).

New methodologies are needed to efficiently investigate the potential for predisposing variants outside of well-established cancer risk genes. Historically, pediatric CNS tumors must have been almost universally fatal causing any germline event associated with high risk of CNS tumors in childhood to be evolutionarily disadvantageous and to likely die out from natural selection. Consequently, genes exhibiting evolutionary intolerance of predicted loss-of-function (pLoF) alterations may serve as areas of particular interest when investigating inherited pediatric cancer susceptibility. A recent study on 141.456 individuals has provided empirical evidence of such highly constrained genes defined by a low LoF observed/expected upper bound fraction (LOEUF) indicating depletion of pLoF variation^8^. The potential of LOEUF score as a marker for evolutionary constraint for the identification of new childhood cancer predisposition genes remains unexplored.

In this nationwide germline WGS study, we seek to establish the prevalence of both pathogenic SNVs and structural variants (SVs) across known cancer predisposition genes in a population-based cohort of 128 children consecutively diagnosed with CNS tumors. Moreover, we hypothesize that pediatric-onset CPS (pCPS) genes show significantly higher constraint than other genes, including adult-onset CPS (aCPS) genes (hypothesis 1). If confirmed, germline pLoF variants in highly constrained genes identified in pediatric cancer cohorts are more likely to be pathogenic than those found in non-constrained genes (hypothesis 2). As a part of the study, these hypotheses are tested and employed to identify novel putative pCPS genes. Lastly, we examine the potential value of systematic pedigree analysis in detecting putatively pathogenic germline variants.

## Methods

### Cohort and sequencing

All children (<18 years of age) diagnosed with primary cancer in Denmark were prospectively offered inclusion over a five-year-period and stratified in a CNS and non-CNS cohort according to primary disease location. As described elsewhere^9^, WGS of leukocyte DNA was performed for each patient and detailed pedigree and medical history information was recorded (detailed in Supplementary Methods).

### Gene panel analysis

SNVs and SVs in a panel of 315 selected cancer related genes^3,10^ were extracted from WGS data and classified by a multidisciplinary team in accordance with ACMG guidelines^11^ (detailed in Supplementary Table 1 and Supplementary Methods).

**Table 1.**
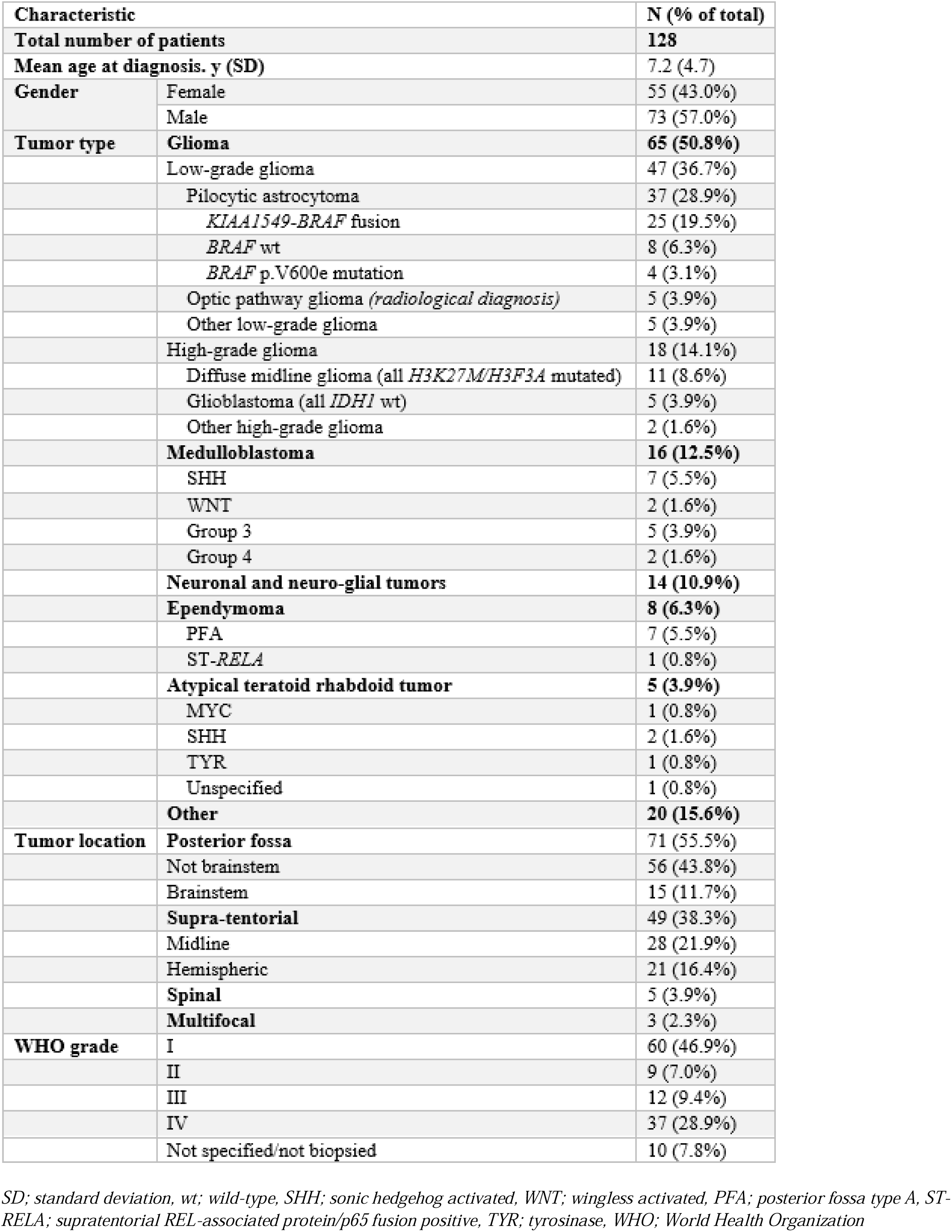
Baseline characteristics.

### Broader gene analyses

Predicted loss-of-function (pLoF) SNVs and SVs were explored in two broader analyses (both detailed in Supplementary Methods):

- Variant burden analysis: The number of pLoF variants in all genes was counted for the CNS and the non-CNS cohorts. Higher pLoF variant burden in the CNS cohort was ascribed to any gene with a rate ratio of three or higher compared to children with non-CNS cancer.
- Constraint gene analysis: pLoF variants among 2,971 evolutionarily constrained genes were extracted and manually curated. Constrained genes with pLoF variants found in the CNS cohort were assessed by scientific literature review and the Gene Ontology (GO) knowledgebase^12^ and String-db^13^.

### Tumor sample investigations

Tumor samples underwent routine histopathological examination including methylation profiling and investigations of DNA mutations and RNA fusions common to the pediatric neuro-oncological population (detailed in the Supplementary Methods).

### Ethical considerations

This study was approved by the Capitol Region Committee on Health Research Ethics (H-15016782) and the Danish Data Protection Agency (RH-2016-219). Oral and written informed consent was collected from all participants and parents/legal guardians depending on age.

### Statistical analysis

Statistical analyses were conducted using IBM SPSS Statistics (v.25) and R (v.3.6.1). The statistical tests used are specified.

## Results

### Baseline characteristics

128 children with CNS tumors (females 43.0%) were included corresponding to an inclusion rate of 84.2% of eligible patients. The main reason for exclusion was language barriers. Median age at diagnosis was 7.0 years (SD 4.7). Gender ratio, tumor type distribution and location (Table 1 & Supplementary Figure 2) were in line with existing population-based reports^1^.

**Figure 1.**
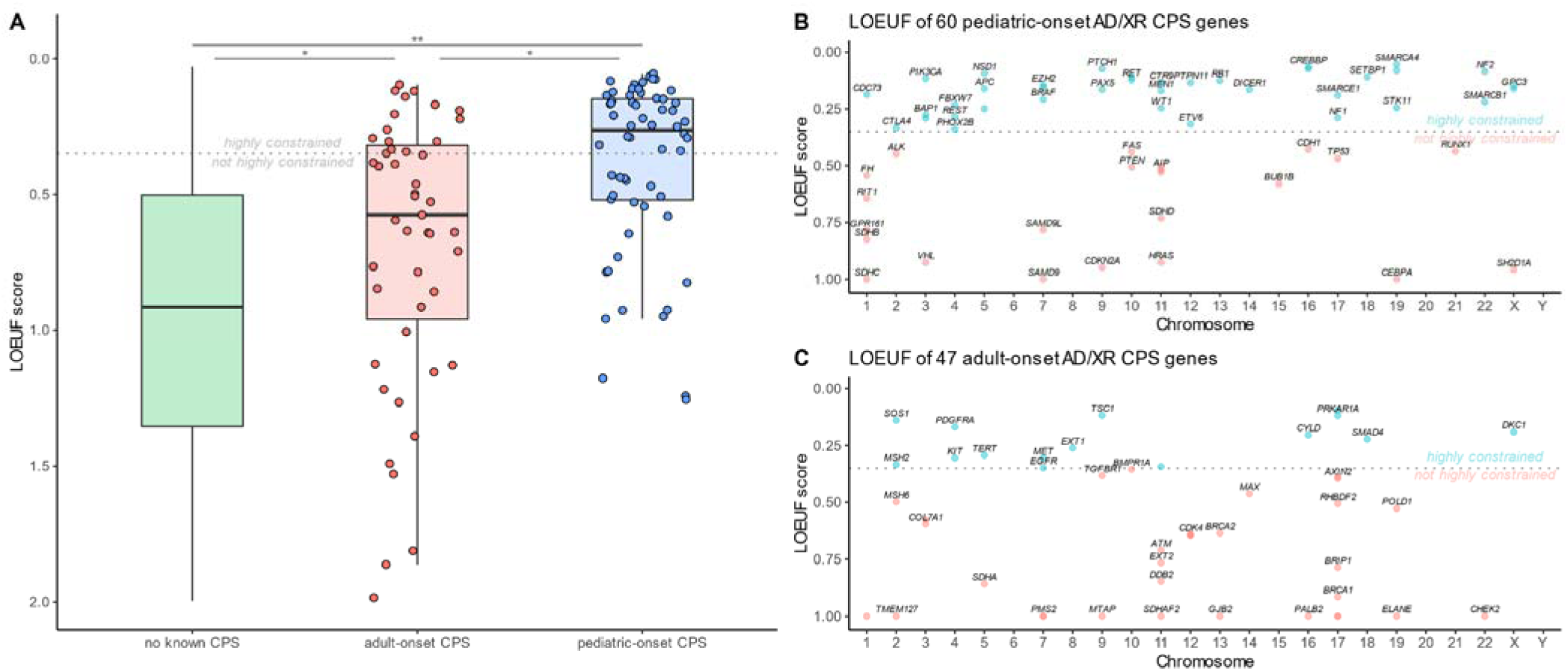
Comparisons of constraint (as determined by LoF variant observed vs. expected upper fraction (LOUEF) score) between genes known to be associated with adult and pediatric cancer risk vs. genes not associated with cancer. **A:** Boxplot comparing LOUEF scores of genes not known to be associated with cancer risk (in green) to adult- and pediatric-onset cancer predisposition syndrome associated genes (aCPS and pCPS in red and blue, respectively). Overlayed jitter plot shows exact distribution of LOEUF scores for aCPS and pCPS genes.* p = 5e^−4^, ** p = 2e^−17^. **B:** Shows genes associated with pCPS for each chromosome and their LOEUF scores, labeled with gene name where possible. The y axis is reversed to show higher constraint higher on the axis. **C:** Same as plot B for genes associated with aCPS. Grey dotted line at 0.35 shows cut-off for high constraint in all panels. Only autosomal dominant and X-linked recessive cancer predispositio syndrome phenotype have been included. In panels **B** & **C** genes with a LOEUF score higher than 1 have been set to 1.00.

**Figure 2.**
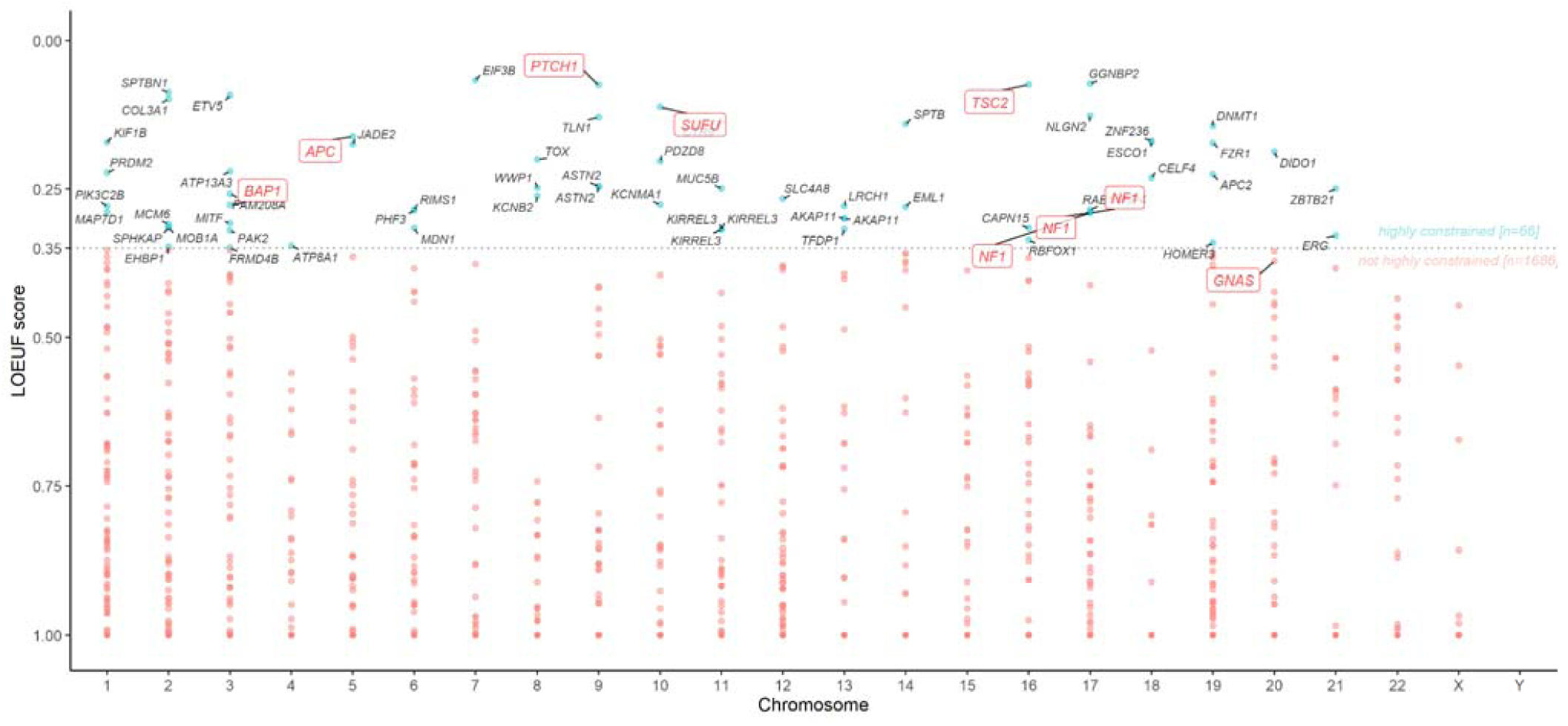
Illustration of all rare predicted loss-of-function (pLoF) variants observed in whole genome sequencing (WGS) data from our cohort. The y axis is reversed to show higher constraint further up on the axis. Genes known to be associated with pediatric-onset cancer predisposition syndromes (pCPS) found on panel analysis are labeled with gene names in red. All genes that showed high constraint (LoF variant observed vs. expected upper fraction (LOUEF) score lower than 0.35) are shown with turquoise dots and labeled with gene names in black. All genes with low constraint (LOUEF score lower than 0.35) are shown with unlabeled red dots. Grey dotted line at 0.35 shows cut-off for high constraint.

### Known cancer related gene findings

WGS data from all 128 patients identified 2,751 SNVs and 985 candidate SVs in the 315 cancer related genes. 13 patients (10.2%) were found to carry pathogenic germline variants (11 SNVs, two SVs). Five *NF1* variants were detected, while the remaining were identified in *APC, BAP1, GNAS, POLE, PTCH1, SUFU, TP53* and *TSC2*. Detailed information on the identified germline variants and relevant clinical data for affected patients are available in Table 2 and 3, respectively. Identical frameshift mutations in *PMS2* [c.2186_2187delTC, p.Leu729Glnfs*6] were identified in two children with pilocytic astrocytoma with *KIAA1549*-*BRAF* fusions. Both were, however, subsequently identified as pseudogene variants by Long Range PCR.

**Table 2.**
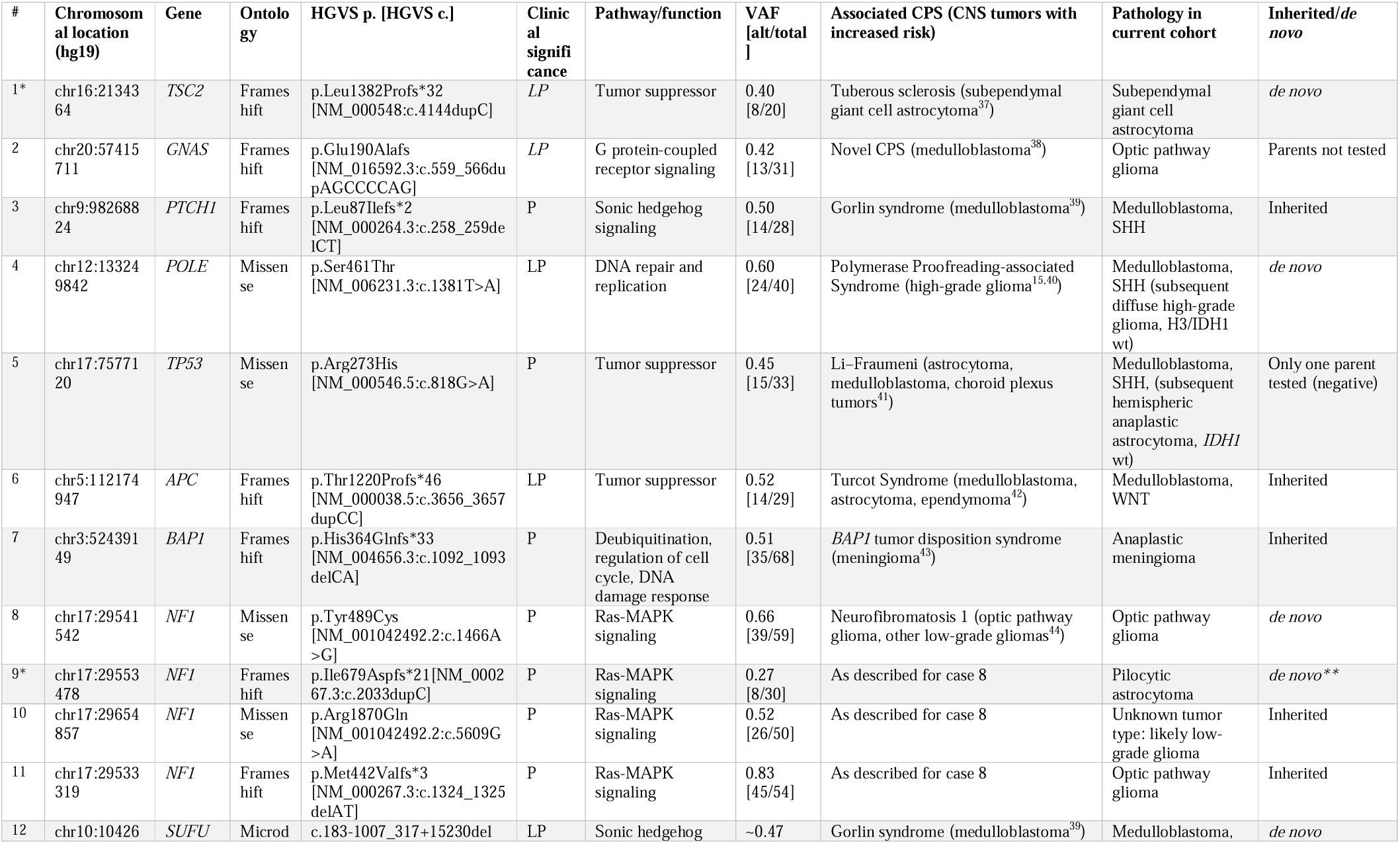
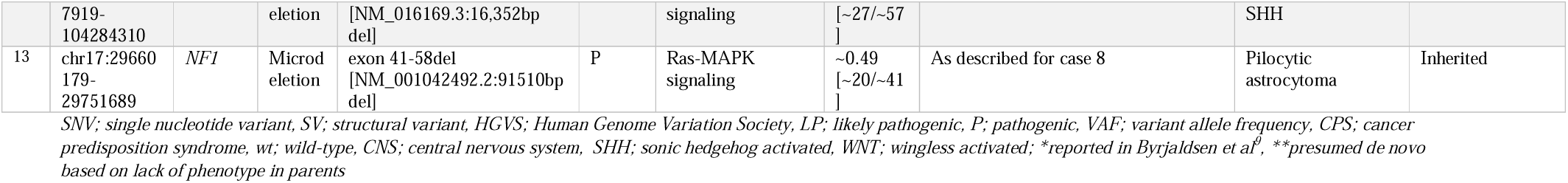
Overview of rare pathogenic variants in known cancer predisposition genes among 128 children with CNS tumors.

**Table 3.**
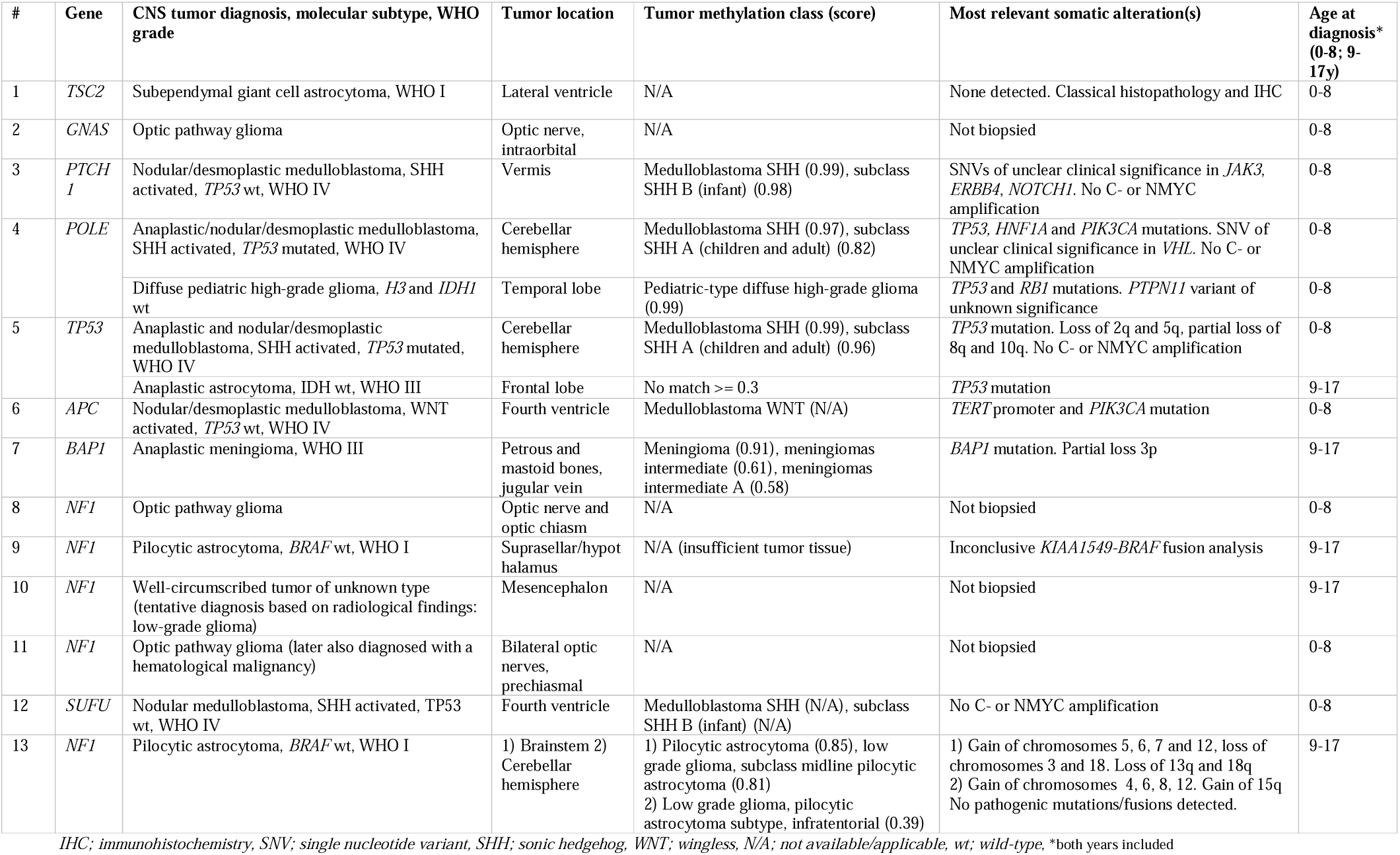
Clinical information for the 13 patients identified with rare pathogenic cancer predisposition gene alterations.

More females tended to harbor a pathogenic CPS gene variant (9/55) compared to males (4/73) (Fisher’s exact test, p=0.073). A lower median age at diagnosis for children with pathogenic variants (4.4 years (SD 5.4) vs. 7.2 years (SD 4.6)) was observed (Mann-Whitney U test, p=0.496). No significant association between major tumor types (Supplementary Table 2) and being affected by a pathogenic CPS gene variant was detected (Fisher’s test, p=0.076).

The tumor type with the highest proportion of patients with pathogenic germline findings was medulloblastoma (5/16), significantly higher than for all other tumor types (8/112) (OR 5.9, CI 1.6-21.2). As expected, the majority of pathogenic variants was found in patients with sonic hedgehog activated medulloblastoma (MB_SHH_, 4/5). The difference in pathogenic germline variant carrier frequencies across medulloblastoma molecular subtypes was not significant (Fisher’s test, p=0.175) (Supplementary Table 3).

Gliomas accounted for just over half of the cohort (50.8%), of which low-grade gliomas made up the majority (47/65). No convincing difference in proportions of pathogenic germline mutations was seen when comparing children with low- and high-grade gliomas (6/47 vs. 0/18, Fisher’s test, p=0.175) or low (I-II, 3/69) and high (III-IV, 6/49) WHO grade tumors (Fisher’s test, p=0.160).

Children with or without a predisposing germline variant did not have significantly different tumor location, defined as supratentorial, posterior fossa, and intraspinal (4/44 vs. 8/68 vs. 0/5, Fisher’s test, p=0.863).

Two children were diagnosed with a second primary CNS tumor during the course of this study: a diffuse high-grade hemispheric glioma, *H3*/*IDH1* wild-type in a child harboring a pathogenic *POLE* variant several years following the primary MB_SHH_ diagnosis (case 4) and a supratentorial anaplastic astrocytoma, *IDH1* wild-type in a child carrying a predisposing *TP53* variant formerly diagnosed with MB_SHH_ (case 5).

Moreover, one patient with an *NF1* frameshift variant diagnosed with bilateral optic pathway glioma also suffered from a hematological malignancy (case 11). The likelihood of being diagnosed with multiple malignancies was significantly higher for carriers of CPS gene variants (3/13 vs 0/115, Fisher’s test, p=8e-4 (p=0.01 when restricted to second CNS tumors)).

### Whole genome variant burden analysis

Burden analysis revealed enrichment of pLoF SVs or SNVs in a myriad of genes in the CNS cohort compared to non-CNS cancer controls (Supplementary Figure 1 & 2). As expected, all nine of the pLoF variants in genes known to cause pCPS were found to be enriched in the CNS cohort. However, variants in a total of 1,533 genes (mean 12.1 per patient) occurred more frequently among cases than controls. Hence, the seven known pCPS genes only constituted 0.5% of all genes identified as enriched. Clearly, burden analysis is of limited use in cohorts with a size and heterogeneity like ours, so we considered whether gene constraint may be more precise in identifying known, and novel, pCPS genes.

### Hypothesis 1: Genes associated with pCPS show significantly higher constraint than both aCPS genes and all other genes

To test Hypothesis 1, a clinical panel of genes associated with pCPS^14^ was compared to a panel of genes associated with CPS regardless of onset^10^. This yielded 60 genes associated primarily with pCPS, while another 47 genes were primarily associated with aCPS. The remaining 19,090 genes were grouped as ‘other’. The three groups showed significant differences in LOEUF scores (Kruskal-Wallis test, p=1e^−19^) and exhibited pairwise significantly lower LOEUF for pCPS genes than for both aCPS related genes (median 0.26 vs. 0.58; Wilcox test, p=5e^−4^) and all other genes (median 0.26 vs. 0.92; Wilcox test, p=2e^−17^) (Figure 1). The seven established CPS genes, in which pathogenic pLoF variants were found in our cohort showed the same trend (mean LOEUF 0.19 vs. 0.95; t-test, p=1e^−4^).

### Hypothesis 2: Germline pLoF variants in highly constrained genes identified in pediatric cancer cohorts are more likely to be pathogenic than those found in non-constrained genes

CNS cohort WGS data harbored 2,149 germline pLoF variants (1,458 SNVs and 691 SVs) in 1,870 distinct genes (Supplementary Figure 1 & 2), of which just 0.4% were known to be associated with pCPSs. Filtering to highly constrained genes, 104 variants across 94 genes in 66 individuals remained (Supplementary Table 4). Of these, manual curation identified 66 (63%) variants in 60 genes as both likely true (high-confidence) and rare among 47 patients. Encouragingly, eight of the nine (89%) pLoF variants in our cohort known to cause pCPSs were found among the 66 variants. Thus, 12% of pLoF variants found in the constrained gene analysis could immediately be appreciated as pathogenic (Figure 2). When subgrouping degree of constraint into deciles, the first, second, third and fourth most constrained deciles of genes had the highest (2/8; 25%), second highest (2/13; 15%), third highest (2/23; 9%) and fourth highest (1/48; 2%) proportion of known CPS genes (two-sided Cochran-Armitage trend test, p=0.021) (Figure 3).

**Figure 3.**
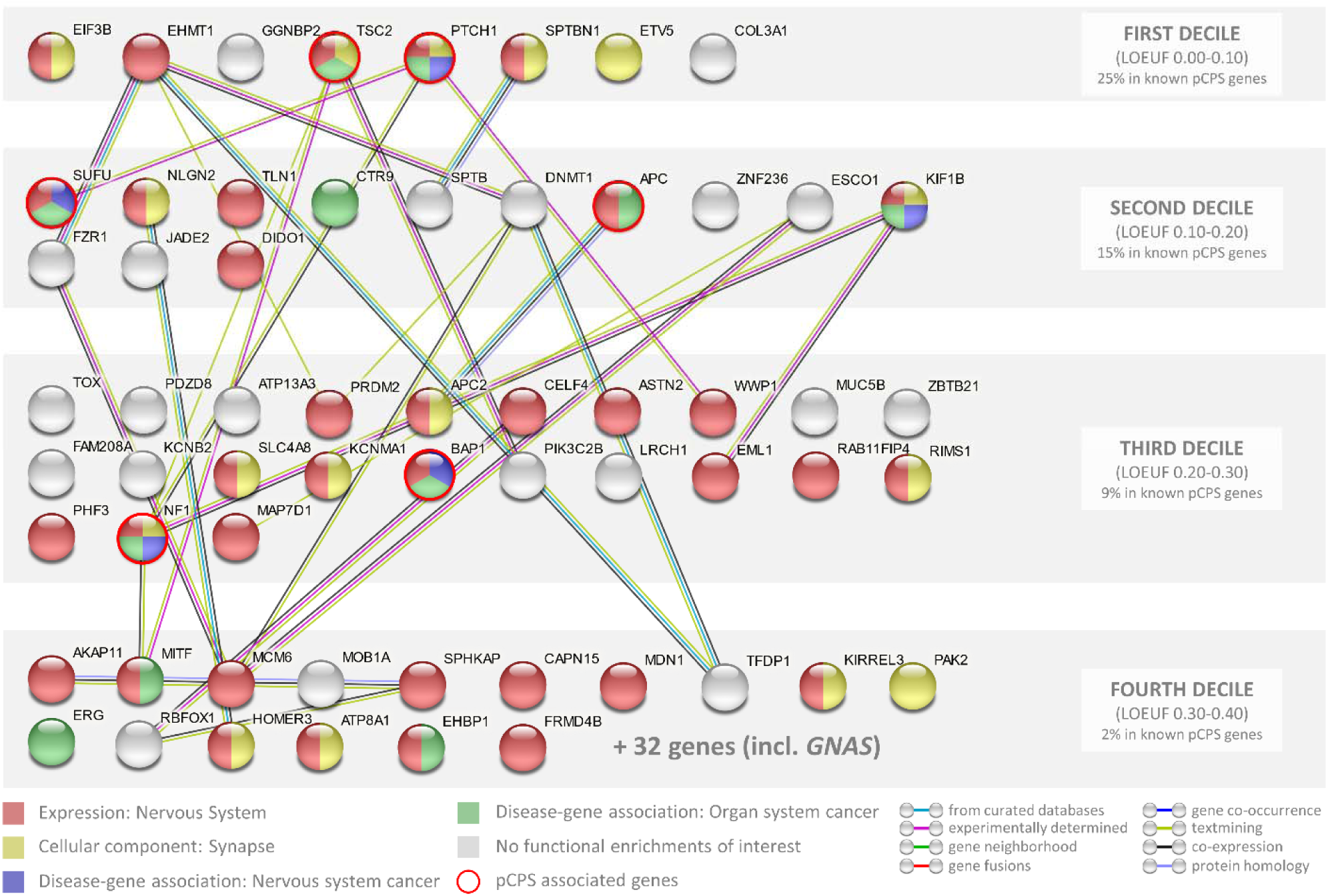
Illustration of the 60 genes found to have predicted loss-of-function (pLoF) variants in constrained genes in our cohort. Genes are ordered from lower (left) to higher (right) LoF variant observed vs. expected upper fraction (LOUEF) score and grouped by decile. Lines illustrate interactions (medium confidence or higher) with color indicating type of interaction evidence based on String-db’s internal algorithm.

The two most constrained gene variants were detected in a child with an anaplastic MB_SHH A_ *TP53* wt, without C-/NMYC amplification and included a heterozygous *EHMT1* 35.5kb deletion (chr9:140592043-140627560) and a heterozygous *EIF3B* frameshift variant (p.Ser590Valfs*12). Several years prior to the tumor diagnosis, the patient had been referred to genetic counseling with mild facial dysmorphia and small biometrics (height, weight and head circumference). Here, the *EHMT1* deletion was identified by microarray in both the patient and one reportedly unaffected parent. SNV and SV analyses of tumor WGS data did not show loss of heterozygosity.

Collectively for all 60 constrained genes, the Gene Ontology (GO) knowledgebase^12^ and String-db^13^ revealed multiple significant enrichments. However, after comparing with the enrichments already present among the 2,960 constrained genes only neuron to neuron synapse cellular component enrichment remained significant (6.34-fold enrichment vs. all genes; FDR=3.7e^−02^. OR 2.28 vs. constrained genes; Fisher’s exact test p = 0.048) (Figure 3).

### Pedigree analysis

3,543 1^st^ to 3^rd^ degree relatives were included in the analysis of pedigrees (available for 122 patients). The mean number registered per family was 29.0 (SD 7.3). No significant differences were seen in the number of 1^st^-3^rd^ degree relatives affected by cancer between families of probands with or without predisposing variants in known CPS genes (3.0 vs 3.7, independent samples T-test p=0.446). Taking into account both the number of relatives with and without cancer and their degree of relation by using the pedigree-based weighted family cancer incidence score did not result in any significant difference (mean score 0.094 vs 0.101, Wilcoxon rank sum test, p=0.648). Limiting the analyses to 1^st^⍰2^nd^ degree relatives, cancers with early onset (<45 years) and neoplasms of the CNS yielded similar inconclusive results (Supplementary Results). Lastly, scores for patients carrying pLoF alterations in constrained genes did not differ significantly from patients without such variants (Wilcoxon rank sum test, 0.092 vs. 0.104, p= 0.318).

## Discussion

In this population-based study, we performed germline WGS of children with CNS tumors to assess the true frequencies and characteristics of pathogenic variants across known CPS genes. In addition, the degree of evolutionary LoF variation intolerance in known pCPS genes was investigated and compared to that of aCPS genes and all other genes. We also illustrate how constrained gene analysis may aid in identifying novel potential pediatric CNS cancer predisposition genes. To our knowledge, this is the first investigation to include constrained gene analysis within pediatric cancer.

### Known cancer predisposition genes

Our findings indicate that ~10% of children with CNS tumors harbor an underlying predisposing variant in a known CPS gene and that such rare, high-risk variant mediated tumor susceptibility varies greatly between tumor types. This is in line with findings from large-scale pan childhood cancer studies using similar cancer gene sets2,3. The detected carrier frequency is significantly lower than the 35% reported by Kline et al4, likely due to their larger fraction of high-grade and recurrent tumors and less stringent variant classification.

Medulloblastoma represented the tumor type with the highest proportion of risk variants within known CPS genes (31%; 5/16). This significantly exceeds the 11% reported by Wasznak et al^6^ in a study on medulloblastoma of all ages. The discrepancy is likely due to an overrepresentation of the MB_SHH_ (44% vs 20%) in our relatively smaller cohort. Our findings clearly support the recent recommendation to offer genetic testing and counseling for children diagnosed with MB_SHH_^6^.

Glioma constituted the most frequent tumor type. Six (9%) children with glioma were found to carry a CPS gene alteration, compared to the 11% reported in a recent WES population-based study including 280 children with astrocytoma^5^. While all detected CPS gene variants in our cohort were found in patients with low-grade glioma, the highest proportion reported by Muskens et al^5^ was among children with glioblastoma. This difference is likely a result of oversampling of high-grade tumors and a larger sample size in the comparator study.

### Novel links between specific tumor entities and established CPS genes

The majority of the observed pathogenic rare CPS gene variants and their associated increased risk of specific brain and spinal cord tumors in children are well-established, e.g. *APC* and MB_WNT_ (Table 2). However, we also detected variants in three such CPS genes not previously linked to the pediatric CNS tumor phenotype found in our study. These included a *GNAS* frameshift mutation in a child with an optic pathway glioma, an inherited *BAP1* mutation in a teenager with an anaplastic meningioma and a *de novo POLE* missense mutation in a patient with MB_SHH_ (Table 2 & 3). Detailed reviews of these cases are provided in Supplementary Discussion. The latter has recently been described in more detail in an independent case series on children with *POLE* variants and constitutional mismatch repair deficiency (CMMRD) syndrome-like phenotypes^15^.

### Constrained gene and variant burden analyses

The pathogenic germline alterations found in 10% of children with CNS tumors were identified through subsetting WGS data to a panel. This revealed 3,736 rare variants of which 13 (0.4%) were found to be pathogenic after careful variant board consideration. Limiting analysis to a panel leads to an underestimation even of the genetic risk identifiable by WGS, as only established CPS genes are assessed. With an estimated 20,000 human genes, it is imperative to develop efficient approaches to focus bioanalytical efforts when investigating WGS data for predisposing variants outside of such known cancer genes. Variant burden analysis is one such approach. Yet, in our heterogeneous data, the known CPS genes constituted only 0.5% of genes with higher variant burden in the CNS cohort.

We formulated and tested a novel approach to gene-disease discovery in pediatric oncology. Genetic predisposition to childhood cancer is necessarily evolutionarily distinct from that of adult malignancies, as variants with high risk of fatal childhood cancers would continuously have been eliminated by natural selection. Recently, vast progress has been made within aggregation of NGS data enabling the identification of pLoF intolerant genes across the human genome^8^. In this study, we find supporting evidence for our Hypothesis 1 stating that genes known to be strongly associated with childhood cancer predisposition show higher pLoF constraint than both adult-onset cancer predisposition genes and other genes in general. In fact, the median LOEUF score for genes associated with pediatric-onset malignancies was shown to be less than half of that of adult cancer predisposition genes and less than a third compared to all other genes. This novel and biologically based method of filtering NGS data to genes exhibiting pLoF constraint thus provides a mechanism of focusing on genomic areas of particular interest to pediatric cancer research - and a potential approach to further uncover heritability of childhood CNS tumors.

Our Hypothesis 2, stating that constraint may identify novel CPS genes, will need further validation in independent pediatric cancer cohorts. However, multiple aspects of our findings support the proposed methodology. Eight out of nine (89%) pLoF variants found among the six genes known to cause pCPS were observed among 60 (10%) genes found in our constrained gene analysis. Additionally, a dose-response trend was observed with larger proportions of known CPS genes within deciles of higher constraint (Figure 3). This raises the question of whether one or more of the remaining 54 genes play a role in CNS tumor predisposition.

We show that these genes tend to be highly expressed in the CNS and are significantly more involved in neuron-to-neuron cellular components than would be expected even within constrained genes. Mounting evidence indicates that neuronal activity plays a critical role in cancer progression, especially in CNS tumors^16^. Somatically, altered neuronal activity has been shown to drive growth of CNS malignancies both through growth factors and through electrochemical synaptic signalling^17^. To our knowledge, this concept has not been described with regard to germline predisposition and our results may inform further research herein.

A heterozygous, inherited deletion within the extremely constrained *EHMT1* gene was detected in a patient with MB_SHH A_. Loss of *EHMT1* causes hypomethylation of H3K9 and this process plays a key role in the pathogenesis of medulloblastoma^18^. Homozygous somatic deletions of *EHMT1* have previously been detected in a molecular study of 1,000 medulloblastomas in two patients; both with the SHH subtype^19^. These somatic deletions were not found in matched germline DNA. Loss of heterozygosity was not detected in the tumor of our patient. Heterozygous germline mutations in *EHMT1* are known to cause Kleefstra Syndrome, which is characterized by intellectual disability, autistic-like features, childhood hypotonia, and distinctive facial features^20^. However, pathogenic/truncating alterations causing Kleefstra Syndrome converge within/prior to the SET-domain located late in the gene (Supplementary Figure 4)^21^, while the deletion in our cohort removes exon 2-4. Deletions inside or across the *EHMT1* gene are absent in more than 10.000 individuals in gnomAD (SV v.2.1). As described, our patient showed a syndromic phenotype extending beyond the cancer diagnosis. Speculatively, early gene deletion may alter, but not eliminate gene function, leading to a phenotype distinct from classic Kleefstra syndrome and perhaps predispose to MB_SHH_.

Other identified constrained genes of apparent interest include, but are not limited to *ASTN2, KIF1B* and *PHF3*. Two patients with medulloblastoma (MB_SHH_ & MB_Grp3_) harbored deletions in *ASTN2*, which encoded protein functions in neuronal migration^22^. *ASTN2* is highly expressed in the cerebellum, including in early cerebellar progenitor cells, from which both MB_SHH_ (migrating granule cell progenitors) and MB_Grp3_ (undifferentiated progenitor-like cells) are believed to originate ^23–25^. Interestingly, *ASTN2* has been shown to be significantly down-regulated in MB_SHH_ with a −3.1 fold change in gene expression compared to non-SHH activated medulloblastoma^26^.

*KIF1B*, in which a pLoF variant was detected in a child with *TP53* mutated MB_SHH_, is highly expressed in fetal cerebellar tissue^27,28^ and has been suggested to act as a haploinsufficient tumor suppressor involved in the pathogenesis of embryonal nervous system tumors such as neuroblastoma, paraganglioma and medulloblastoma^29–31^. The patient also carried the described pathogenic missense variant in the *POLE* gene. Of interest, a child with di-genic *POLE* and *PMS2* pathogenic variants and MB_SHH_ was recently reported, suggesting that cancer predisposition driven by germline *POLE* variants may have important modifiers^32^.

Another pLoF variant was detected in *PHF3* in a patient with a midline glioblastoma, *IDH* wt. Interestingly, downregulation of *PHF3*, which has been shown to occur frequently in glioblastoma^33^, has recently been suggested to drive glioblastoma development by depression of transcription factors that regulate neuronal differentiation^34(p3)^.

### Pedigree analysis

Family cancer incidence did not differ significantly between children with or without predisposing germline alterations, which is in line with findings in comparable cohorts^3,35^. The introduced novel pedigree-based family cancer incidence score, which weighs both the number of relatives registered with and without cancer and their relation to the proband, also did not differ between families of probands harboring pathogenic CPS gene variants. Consequently, our data does not support family history as a sole indication for genetic testing.

A high family cancer incidence would be expected to result from inherited highly penetrant variants. The limited predictive power of pedigrees possibly reflects that variants associated with high childhood cancer risk tend to be *de novo* and/or located in highly constrained genes. While variants with moderate or low penetrance may not infer sufficient risk to create a detectable cancer signal in pedigrees. Our sample size limited stratification by *de novo* status.

### Strengths and weaknesses

Key strengths of this study include; a prospective population-based design (Supplementary Figure 4) and a combination of WGS data and deep phenotyping, up-to-date neuropathology reports including methylation profiling and detailed clinical data and multigenerational family histories. Also, our study included SV detection and went beyond panel-based analysis, the value of which is illustrated by the pathogenic *SUFU* and *NF1* deletions detected and by findings from the burden and constrained gene analyses.

The relatively short and variable length of follow-up made investigations into correlations between germline variants and prognosis/survival unjustified. Meaningful comparisons of age of onset and pedigree-based incidence scores for children harboring pLoF variants in constrained genes other than known pCPS genes were limited by sample size. Moreover, parental sequencing was only available for cases with pathogenic alterations in known CPS genes - not other constrained genes.

However, as the cohort will continue to increase in size and length of follow-up, assessment of the role of germline variants for treatment response, toxicity and patient outcomes will become possible. Optimally, a large whole-genome sequenced control cohort of healthy, ethnically comparable children will be available for such future investigations. This was not the case for the current study, and the use of a pediatric non-CNS cancer cohort may have affected our burden analysis in a conservative direction. The main reason for exclusion was lack of Danish or English language proficiency which may have conferred exclusion bias towards certain ethnical minorities. Restricting inclusion of children with optic pathway gliomas to patients who received active treatment may have negatively affected the cohort prevalence of *NF1* variants. SV analyses included only deletions detectable on WGS, which, while generally superior to panel or WES, identifies fewer SVs than third generation sequencing^36^.

*In summary*, this population-based study establishes that ~10% of pediatric brain and spinal cord tumors can be attributed to rare variants in known CPS genes. Moreover, we introduce a novel approach to investigate pLoF variants in constrained genes and how this methodology may increase the understanding of genetic susceptibility in children with CNS tumors. Our findings clearly illustrate the importance of assessing both SVs and SNVs when investigating genetic predisposition to childhood cancer. These results have direct implications for clinical genetic counseling, may inform future novel gene-disease association studies and add to the mounting evidence of genetic predisposition in pediatric neuro-oncology.

## Data Availability

All data produced in the present work are contained in the manuscript with the exception of genetic sequencing data. Danish legal regulation does not permit uploading of raw sequencing data. Selected data may be made available upon reasonable request (dependent on required approvals from relevant scientific ethic boards) to the authors

