## Supplementary Material for "Population-based whole-genome sequencing with constrained gene analysis identifies predisposing germline variants in children with central nervous system tumors"

#### Supplementary Methods

*Cohort and sequencing*

All children (<18 years of age) diagnosed with primary cancer, including CNS tumors, have since 2016 been offered inclusion in the "Sequencing Tumor And Germline DNA—Implications for National Guidelines (STAGING)" study[1]. Participation covers but is not limited to i) WGS of germline DNA, ii) targeted sequencing of parental DNA for cases with detected predisposing variants in known cancer genes, iii) collection of a pedigree and detailed medical history and iv) genetic counseling. The national study setup and protocols for inclusion have previously been described in detail[1]. The current study covers the pediatric CNS tumor sub cohort of the STAGING study included from July 1st 2016 until July 1st 2021 and encompasses solid histopathologically diagnosed intracranial- and intraspinal tumor entities with the exception of germ cell and secondary tumors. Children with non-biopsied tumors, such as radiologically diagnosed optic pathway gliomas, were included in STAGING if active therapy (i.e. surgery, chemo- or radiotherapy) was initiated.

Leukocyte DNA was isolated from peripheral blood samples drawn alongside standard blood-sampling executed as part of treatment. When possible, parental blood samples were taken to establish whether detected pathogenic variants were inherited or occurred *de novo*. Sequencing protocols have been published in detail elsewhere[1]. In brief, WGS was performed using the HiSeqX platform (Illumina, San Diego, CA, USA) with paired-end sequencing of 150-bp reads and target 30X average coverage. Reads were mapped to the hg19 reference genome sequence (GRCh37.p13; RefSeq assembly accession GCF_000001405.25) using GATK version 3.8 or the DNAseq pipeline (Sentieon, San Jose, CA, USA). VarSeq software (version 2.2.3, Golden Helix, Bozeman, MT, USA) was used to annotate variants. Long-range (LR) polymerase chain reaction (PCR) for assessment of pseudogene status for *PMS2* variant carriers was conducted as previously described[2].

*Collection and analysis of family pedigrees*

Pedigrees covering 1st–3rd degree relatives were constructed through systematic interviews conducted by trained staff with patients and/or their parents or legal guardians. Interviews were scheduled days to weeks in advance to allow for needed preparation time for the participants. 1st degree relatives were defined as parents and siblings, 2nd degree as grandparents, uncles/aunts and half-siblings while 3rd degree relatives covered great grandparents, grandparents' siblings, cousins and half-uncles/-aunts.

Pedigrees were analyzed by summarizing the number of relatives in each degree. In addition, all incidences of cancer including cancer type, age at diagnosis, tobacco use and relation to the proband were registered. A novel pedigree-based weighted family cancer incidence score was calculated based on the following equation:

$$\frac{((n\ 1st\ degree\ relatives\ w/\ cancer \times 0.5)+(n\ 2nd\ degree\ relatives\ w/cancer\ \times 0.25)+(n\ 3rd\ degree\ relatives\ w/cancer \times 0.125))}{((n\ 1st\ degree\ relatives\ \times 0.5)+(n\ 2nd\ degree\ relatives \times 0.25)+(n\ 3rd\ degree\ relatives\ \times 0.125))}$$

*Gene panel analysis*

Resulting sequencing data was filtered for pathogenic SNVs and SVs in a panel of 314 cancer related genes selected from the existing medical literature, 33 of the actionable genes identified by the American College of Medical Genetics and Genomics (ACMG v2.0)[3] not already included in the panel (Supplementary Table 5), and the newly identified medulloblastoma predisposition gene *ELP1*[4] Detected variants were reviewed by a multidisciplinary team of clinical geneticists, pediatric oncologists and bioinformaticians and classified as "pathogenic", "likely pathogenic", "variants of unknown significance", "likely benign" and "benign" in accordance with current international standards[5] In our study, variants classified as "pathogenic" or "likely pathogenic" are referred to as simply "pathogenic".

*Broader gene analyses*

SVs were called for all patients based on aligned WGS data using Manta (1.4), CNVnator (0.3.3), CNV kit (0.9.6) and Delly2 (0.8.1) using R (3.6.1). All children with non-CNS cancers in the STAGING study (n=467) as well as WGS-called SVs from a Danish pediatric clinical non-cancer cohort (n=210, critically ill children under suspicion of metabolic disease or epileptic encephalopathy) served as controls in the SV burden analysis. SVs detected in the non-cancer cohort were removed, as were non-exonic SVs together with inversions and insertions. Remaining SVs were quantified in patients with CNS and non-CNS disease and number of SVs per gene were compared. Enrichment was ascribed to any gene with a higher or equal variant count in CNS. Due to cohort size differences, equal counts amounted to a 3-fold increase rate ratio. Any genes with a higher count of variants in non-CNS than CNS were ascribed non-enriched (Supplementary Figure 1).

Similarly, pediatric non-CNS cancer patients in the STAGING study served as controls for the supplementary SNV analyses. Called SNVs were filtered by removing intronic and non-LoF SNVs and by application of the following quality control (QC) parameters; coverage >15X, VAF >0.3 and <0.70, strand bias <10, allele count =2, indel size <10. SNVs with >2 exact matches among non-CNS cancer patients were also removed (Supplementary Figure 2). SNV burden analysis was performed as for SVs (Supplementary Figure 2).

Subsequently, both the SV and SNV analyses were restricted to pLoF variants in constrained genes defined by a LOEUF </= 0.35 (recommended threshold[6,7]). LOEUF scores were derived from canonical transcripts in Supplementary Dataset 11 in Karczewski et al[6]. Resulting variants underwent manual curation based on visual analysis of WGS data using Integrated Genome Viewer , comparison to The Genome Aggregation Database (gnomAD v2.1)[6] for population frequencies and ClinVar[8] for variant classification as well as scientific literature review.

*Tumor sample investigations*

Tumor samples from all patients who had a biopsy or resection surgery performed underwent routine neuropathological examinations including microscopy, immunohistochemistry, targeted next generation sequencing (NGS) of mutations common to the pediatric neuro-oncological population, multiplex-ligation probe amplification (MLPA) and DNA methylation profiling upon the preference of the pediatric neuropathologist. Methylation profiles were attained using Illumina Human Methylation 450 BeadChip or EPIC BeadChip arrays. Unprocessed IDAT files were uploaded to and processed by the publicly available classifier tool (www.molecularneuropathology.org) to establish tumor methylation class[9].

#### Supplementary Results

*Pedigree analysis limited to 1st-2nd relatives, early-onset cancers and neoplasms of the CNS*

When limiting the analysis to 1st-2nd degree relatives, cancers diagnosed before the age of 45 years or neoplasms of the CNS, the cancer incidence score was marginally higher for families of probands with pathogenic germline variants (mean score 0.269 vs 0.211, 0.077 vs 0.076 and 0.058 vs 0.028, respectively) although these findings were not statistically significant (Wilcoxon rank sum test, p-values 0.350, 0.417 and 0.634). Our sample size limited stratification by *de novo* status and parental sequencing was only available for cases with pathogenic alterations in known cancer genes - not other constrained genes.

*Manual curation of pLoF variants in constrained genes*

All pLoF variants called within constrained genes were manually curated for veracity and possible high-penetrance pathogenicity using the following flags/criteria:

- **Late truncation:** Variants leading to a truncation within the latter 5% of the gene's coding sequence were flagged.
- **Non-disrupting to splice:** Variants found to not disrupt splice were flagged.
- **Splicing impact unknown:** Novel variants within known splice sites where functional studies of this or similar variants have not been performed were flagged.
- **Gnomad frequency:** Variants where either the exact or identically truncating variants were observed in more than one in 5000 alleles were flagged.
- **Low complexity region:** Variants located in low complexity regions such as trinucleotide repeat segments were flagged.
- **Deep intronic:** Variants located more than 100 bp inside an intron were flagged.
- **Low-quality site:** Variants located in areas marked as low-quality, i.e. poor coverage, areas were flagged.
- **Weak PhyloCSF:** Variants in a region that does not exhibit expected conservation patterns expected for a coding exon were flagged.
- **No split reads (SV only):** Whole genome sequencing data did not demonstrate split reads at the breakpoints identified in the called structural deletion.
- **No coverage drop (SV only):** Whole genome sequencing data did not demonstrate drops in coverage in the region called as a structural deletion.

Of the 104 pLoF variants identified in constrained genes, 38 were flagged for one or more of the criteria above. 14 constrained genes called with structural deletions, of these 13 were flagged due to no coverage drop and no observed splits read and one was true but failed to high gnomad frequency. Among SNVs, the following flags were used; deep intronic (3), Gnomad frequency (5), late truncation (3), late truncation, gnomad frequency (1), late truncation and splicing impact unknown (2), low complexity region (8), non-disrupting to splice (1), weak PhyloCSF (1) (Supplementary Table 4).

*Description of cases with rare variants in extremely constrained genes*

The third most constrained event, a heterozygous *GGNBP2* splice variant (NM_024835.4:c.1366+2T>C), was found in a teenager with a hemispheric diffuse astrocytoma, *IDH1* mutated, *BRAF* wt, WHO II. Despite being absent from gnomAD, an identical variant was also identified in a patient in our non-CNS cohort.

The fourth most constrained event, a heterozygous *SPTBN1* 4.5kb structural deletion (chr2:54897246-54901726), was observed in a teenager known with an autism spectrum disorder who suffered from a diffuse intrinsic pontine glioma, *H3K27M* mutated, WHO IV. This patient also harbored a second variant in another (lesser) constrained gene (*ATP8A1*).

The fifth most constrained event, a heterozygous *ETV5* structural deletion (chr3:185780365-185785187), was observed in young child with a hemispheric anaplastic pleomorphic xanthoastrocytoma, *IDH1* wt, *BRAF* [c.1799T>A, p.Val600Glu] mutated, WHO III. As the latter patient, this child also carried a second event in a less constrained gene (*CAPN15*).

#### Supplementary Discussion

*Novel links between specific tumor entities and established cancer predisposition genes*

The majority of the observed cancer predisposition gene variants and their associated increased risk of specific brain and spinal cord tumors in children are well-established (Table 1). However, three known cancer risk gene variants not previously linked to the pediatric CNS tumor entities in question were identified, including a *GNAS* frameshift mutation in a young child with a radiologically diagnosed optic pathway glioma (case 2). While somatic *GNAS* alterations are frequent across a range of cancers[10], germline mutations have only been reported in two cases of pediatric medulloblastoma[11,12] and not previously in a patient with optic pathway glioma. Such variants have also recently been linked to childhood high-penetrance childhood obesity[13]. Prior to treatment, our patient had a weight and BMI both exceeding the 99th percentile and remains obese. The previous case reports have not reported weight.

A *de novo POLE* missense variant was identified in a child with café au lait spots diagnosed with a hypermutated anaplastic/nodular/desmoplastic $MB_{SHH\ A}$, *TP53* mutated, WHO IV (case 4). *POLE* variants have previously only been reported in a limited number of patients with high-grade glioma[14,15] and in a single recent case of a five-year-old child with a hypermutated non-WNT/non-SHH medulloblastoma[16]. The latter patient, in which tumor methylation profiling was not reported, was described with café au lait macule features similar to those of our patient. The *POLE* germline mutation identified in this cohort is thus believed to be the first reported for $MB_{SHH}$ and has recently been described in more detail in an independent case series on children with *POLE* variants and constitutional mismatch repair deficiency (CMMRD) syndrome-like phenotypes[17].

Pathogenic *BAP1* variants have long been known to increase the risk of a number of different non-CNS cancer types[18(p1)]. However, over the last decade, several cases of meningioma in adults with *BAP1* germline variants have been reported[18–20]. This inborn predisposition has mainly been detected in highly aggressive tumors with rhabdoid features similar to that of the anaplastic meningioma (with rhabdoid features), *BAP1* mutated, WHO III, diagnosed in a teenager in our cohort (case 7)[20]. A parent of the proband harbored the same frameshift variant and suffered from a rare form of melanoma, which has only once before been reported as a part of the *BAP1* tumor disposition syndrome[21].

The *SUFU* tumor suppressor gene is well established as a major medulloblastoma predisposition gene[22], in which pathogenic variants may cause Gorlin's syndrome and increase risk of medulloblastoma. *SUFU* deletions, such as the 16kb truncating deletion detected in the young child with a nodular/desmoplastic $MB_{SHH\ B,}$ *TP53* wt, WHO IV in our cohort (case 12), however, are much rarer events and have only been reported in a very limited number of patients with medulloblastoma, both with and without Gorlin's syndrome[23–26].

*Assessment of population coverage*

Most previous studies investigating germline predisposition in childhood cancer using similar methodologies (WGS/WES of germline DNA) include cohorts from highly specialized tertiary pediatric oncology centers. This may infer risk of selection bias towards certain tumor types treated at the given centers (e.g. higher grade tumors, recurrent tumors) and/or patients of different ethnic and socioeconomic backgrounds (depending on geography, culture, the organization of the involved healthcare systems, insurance practices etc). On the contrary, STAGING is a nationwide study which offers inclusion to all children diagnosed with cancer before the age of 18 years in Denmark. All children with CNS tumors in Denmark are treated at one of three specialized public pediatric neuro-oncology centers, while pediatric neuro-oncological surgery is limited to two of these three centers. All involved centers participate in STAGING.

To assess the coverage of the current study's population-based design we conducted a comparative audit using data from the Danish Childhood Cancer Registry (DCCR). The DCCR is a nationwide clinical quality database surveilling various indicators of childhood cancer. The combination of the Danish public healthcare system and the unique Danish personal identity number allows for cross-linkage between the DCCR and other disease registries including the Danish National Patient Registry, the National Pathology Registry and the Danish Cancer Registry thereby strongly limiting the degree of missing data. Consequently, the DCCR reports data completeness for children diagnosed with cancer below 15 years of age in Denmark of 100%[27]

As seen in Supplementary Figure 4, STAGING covered 82% of children diagnosed with a CNS tumor nationwide during the study period according to the DCCR. The percentage of patients not identified by STAGING differs slightly between tumor types. In particular, children in the DCCR listed with non-specified tumors without histology report conclusions appear to have been missed to a higher degree than others. This might reflect that STAGING does not offer inclusion to patients with conservatively managed, non-biopsied optic pathway gliomas. Also, this category specific high degree of missingness may reflect pathologies not considered as CNS tumors by the including/treating clinical staff and/or inconsistent inclusion practices for children with "incidentalomas" and other not actively treated intracranial tumors.

### Supplementary Figures

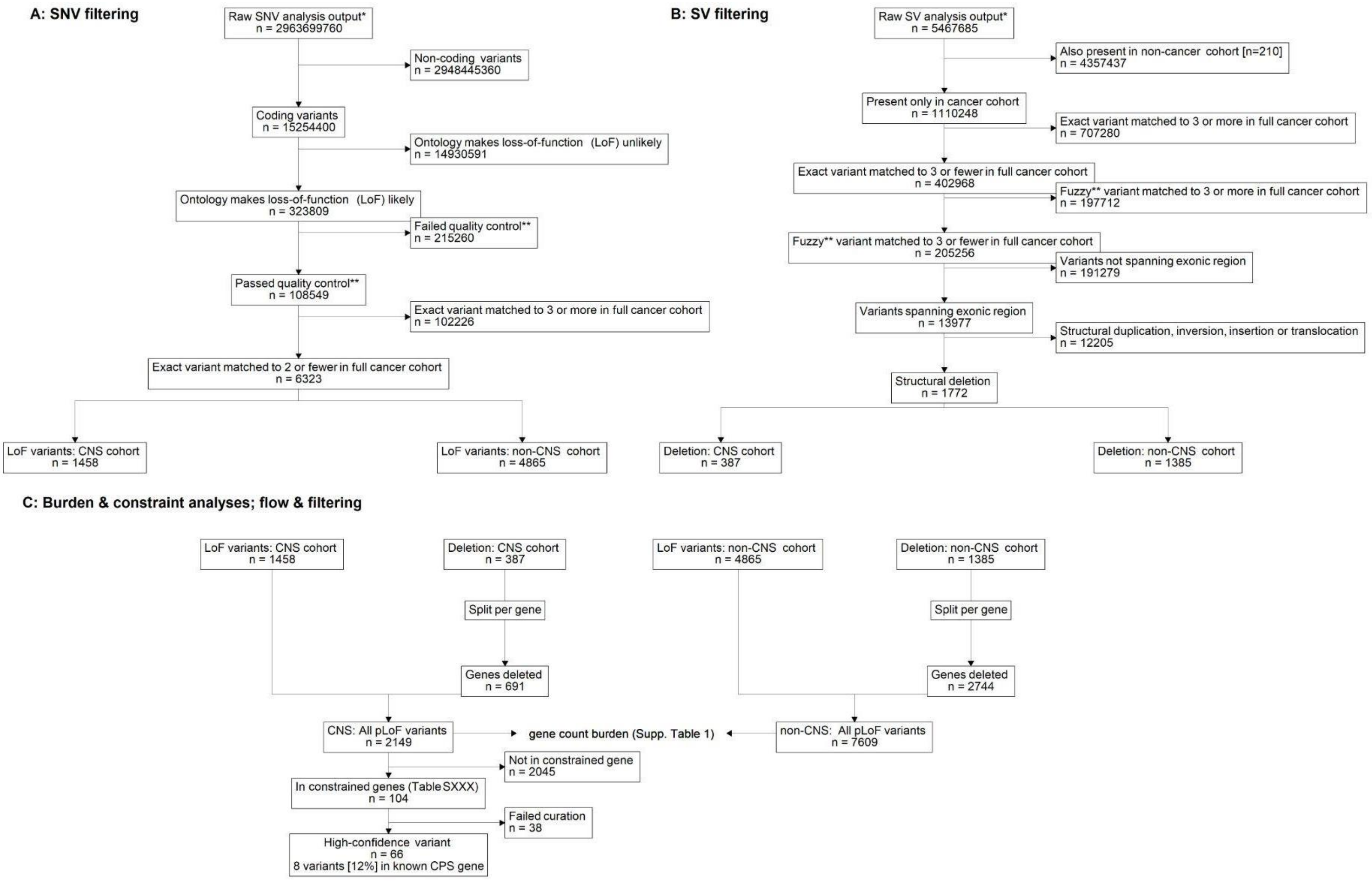


**Supplemental Figure 1: A** and **B**: Consort-style chart quantifying how the raw data output of single nucleotide and structural variant (SNV/SV) calls were filtered.
**C**: Using the post-filtering data from panels above, the flowchart illustrates the combining of deletions split per gene and loss-of-function (LoF) ontology SNVs for both CNS cohort [n =120] and non-CNS [n = 595] for burden analysis (Supplemental Table 1). Finally, the combined LoF variants in the CNS cohort were filtered to constrained genes only [defined by having LOEUF <= 0.35] and manually curated (details in Supplemental Table XXX). * All SV calls are based on Manta ** Fuzzy matching was performed by rounding the chromosome coordinates to -2 decimals.

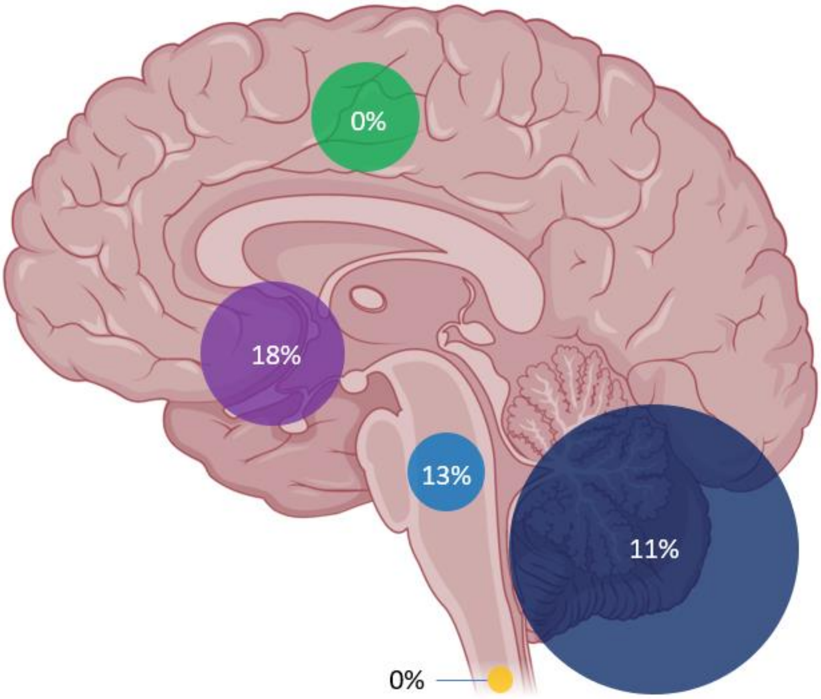


**Supplemental Figure 2:** Circle size is representative of the number of patients with tumors in each location. Assigned percentage annotates the known cancer predisposition gene pathogenic germline variant carrier frequency of the specific tumor location. *Green; supratentorial hemispheric, purple; supratentorial midline, light blue; brainstem, dark blue; cerebellum, yellow; spinal.* Created with BioRender.com

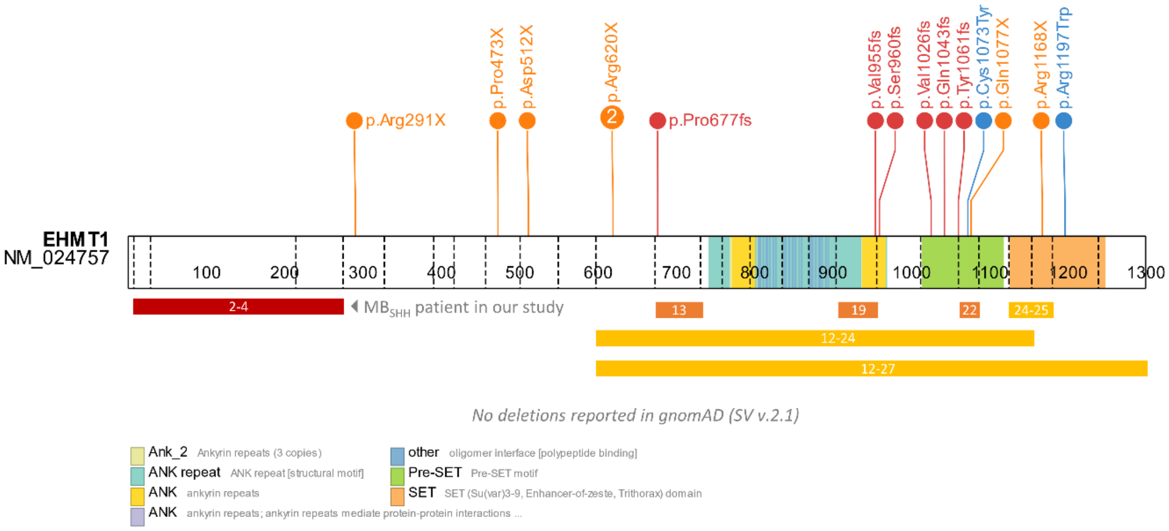


**Supplemental Figure 3:** Shows the protein product of the canonical transcript of the *EHMT1* gene. The red bar below the protein represents the heterozygous deletion found in our patient spanning exons 2-4. Heterozygous pathogenic variants causing Kleefstra Syndrome are represented by lollipop markers showing protein change and colored by ontology (orange: nonsense, red: frameshift, blue: missense) and bars representing exons affected (yellow: deletion, orange: exon skipping).

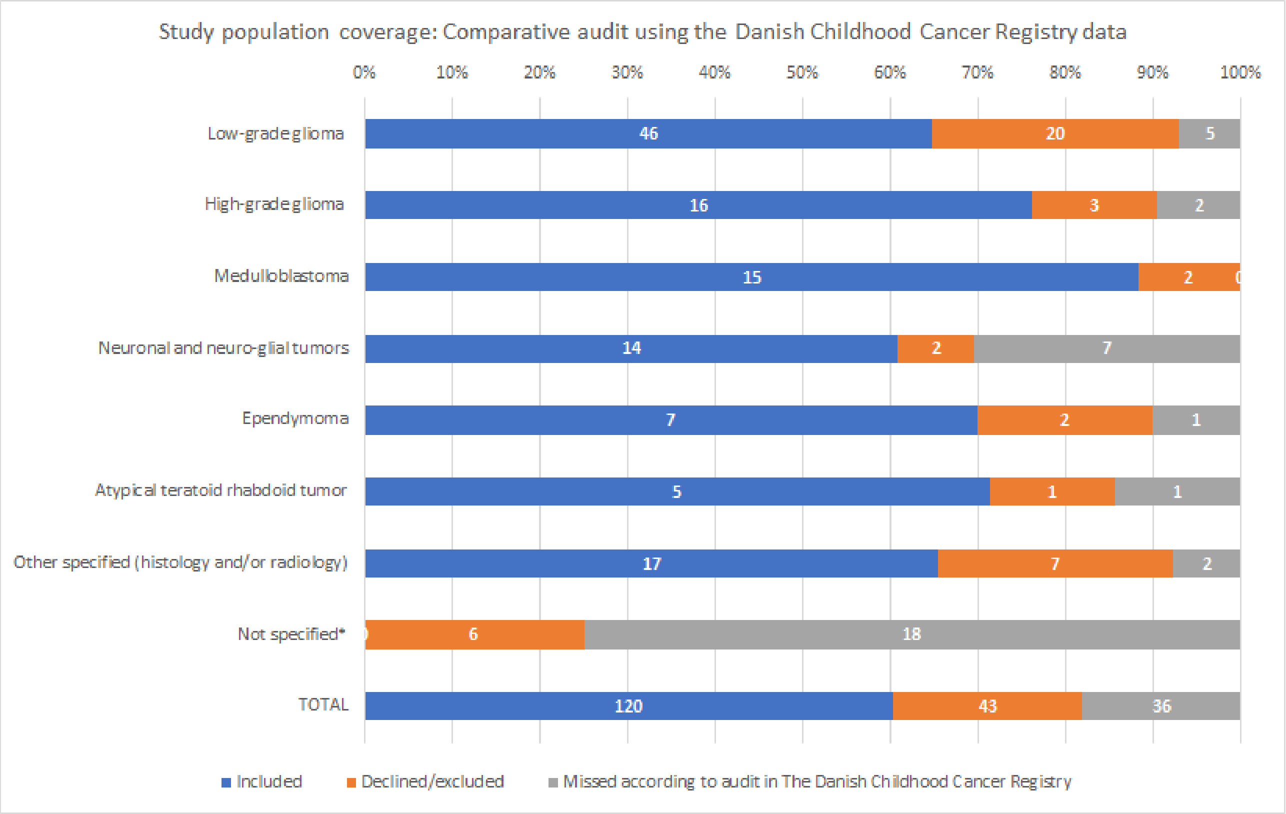


**Supplemental Figure 4**. Illustration of the study's population coverage by comparison with data from the Danish Childhood Cancer Registry for patients diagnosed with CNS tumors before the age of 18 years nationwide. Restricted to diagnoses registered between July 1st 2016 - December 31st 2020 (both dates included) to ensure harmonization with available registry data, which accounts for the discrepancy (n = 8) compared to the full cohort of 128 cases. Data labels represent the number of patients. Germ cell tumors and non-neoplastic cystic lesions of the CNS (e.g. colloid cysts, Rathke's cleft cysts) are not included. *No registered histopathological diagnosis or specific tumor location.

#### Supplementary Tables

**Supplemental Table 5. Variant burden results**

| # of variants in gene in the **CNS cohort** [n =128] | # of variants in gene in the **non-CNS cohort** [n = 467] | **Rate ratio** of variant counts in gene | Selected genes. All **known CPS genes** are listed and in bold. |
|---|---|---|---|
| 4 | 2 | 7.30 | *SULT1A1, ASMT* |
| 3 | 3 | 3.96 | *FLG2, COL6A5, DOCK6* |
| 3 | 2 | 5.94 | *PSG2, TSNARE1, UBXN11, SLCO1B3, DNAH17, HTT* |
| 3 | 1 | 11.88 | *ZAN, DEPDC4, FBN3, CCDC81, TRPM2, ALDH1L2, SLC27A3, ALDH1B1* |
| 3 | 0 | Infinite | ***NF1***, *C9orf135, C6orf99, DHX57, ENO4, SIRT3, ZNF747, AGAP6, KIRREL3* |
| 2 | 2 | 3.96 | 25 genes |
| 2 | 1 | 7.92 | 56 genes |
| 2 | 0 | Infinite | 68 genes |
| 1 | 1 | 3.96 | 420 genes |
| 1 | 0 | Infinite | ***SUFU, TSC2, PTCH1, APC, BAP1, GNAS*** + 921 other genes |
| 0 to 4 | 1 to 13 | < 3.00 | 4,629 genes |
